# Productivity costs associated with reactive school closures related to influenza or influenza-like illness in the United States from 2011 to 2019

**DOI:** 10.1101/2023.01.05.23284221

**Authors:** Joohyun Park, Heesoo Joo, Brian A. Maskery, Nicole Zviedrite, Amra Uzicanin

**Affiliations:** Division of Global Migration and Quarantine, Centers for Disease Control and Prevention, Atlanta, Georgia, United States of America

**Author notes:** **Corresponding author:** Joohyun Park, PhD, Division of Global Migration and Quarantine, Centers for Disease Control and Prevention, 1600 Clifton Rd. NE, MS H16-4, Atlanta, GA 30333, USA. **Disclaimer:** The findings and conclusions in this study are those of the authors and do not necessarily represent the official position of the U.S. Centers for Disease Control and Prevention.

## Abstract

**Introduction:** Schools close in reaction to seasonal influenza outbreaks and, on occasion, pandemic influenza. The unintended costs of reactive school closures associated with influenza or influenza-like illness (ILI) has not been studied previously. We estimated the costs of ILI-related reactive school closures in the United States over eight academic years.

**Methods:** We used prospectively collected data on ILI-related reactive school closures from August 1, 2011 to June 30, 2019 to estimate the costs of the closures, which included productivity costs for parents, teachers, and non-teaching school staff. Productivity cost estimates were evaluated by multiplying the number of days for each closure by the state- and year-specific average hourly or daily wage rates for parents, teachers, and school staff. We subdivided total cost and cost per student estimates by school year, state, and urbanicity of school location.

**Results:** The estimated productivity cost of the closures was $476 million in total during the eight years, with most (90%) of the costs occurring between 2016−2017 and 2018−2019, and in Tennessee (55%) and Kentucky (21%). Among all U.S. public schools, the annual cost per student was much higher in Tennessee ($33) and Kentucky ($19) than any other state ($2.4 in the third highest state) or the national average ($1.2). The cost per student was higher in rural areas ($2.9) or towns ($2.5) than cities ($0.6) or suburbs ($0.5). Locations with higher costs tended to have both more closures and closures with longer durations.

**Conclusions:** In recent years, we found significant heterogeneity in year-to-year costs of ILI-associated reactive school closures. These costs have been greatest in Tennessee and Kentucky and been elevated in rural or town areas relative to cities or suburbs. Our findings might provide evidence to support efforts to reduce the burden of seasonal influenza in these disproportionately impacted states or communities.

## Introduction

Living in a household with a person ill with influenza more than triples the attack rate of the illness for the rest of the household [1]. As children can play a major role in the spread of influenza within households [2], closures and dismissals of child care facilities and K−12 schools have been implemented as a community-level nonpharmaceutical intervention during influenza pandemics to prevent virus transmission [3–5]. Preemptive, coordinated school closures and dismissals are recommended for use early when an influenza pandemic is severe, very severe, or extreme, with a consideration of secondary consequences such as loss of productivity for parents or school employees and loss of access to subsidized school lunches for children [3, 5]. During the coronavirus disease 2019 (COVID-19) pandemic, nationwide school closures were implemented preemptively in the United States; statewide mandates or recommendations were issued for public school closures among 50 states and the District of Columbia (D.C.) between mid- and late-March 2020 [6].

School closures also occur reactively during seasonal influenza epidemics or pandemics in situations where considerable influenza transmission has already occurred in schools and surrounding communities causing high levels of student and staff absenteeism due to illness [3, 7]. Reactive school closures may be short in duration and, because they are implemented late during an outbreak, they are unlikely to be effective in reducing virus transmission in the communities in the United States [3, 8–11]. As any school closures other than planned academic breaks, reactive closures are disruptive for school process and may lead to costs and consequences for households, including parents missing work and losing wages to provide child care or paying for child care. Some families would also lose access to free or subsidized school lunch programs [8, 11–13].

Although there are many published studies on the economic costs of hypothetical, preemptive school closures during simulated influenza pandemics in the United States [14, 15]; the costs of relatively short-term reactive school closures (i.e., usually 1- or 2-day closures) have not been previously reported. This analysis provides an estimate of these costs by using previous surveys of the socioeconomic consequences of reactive school closures associated with influenza or influenza-like illness (ILI) [11]. This analysis aims to estimate the costs in terms of lost productivity for school staff and for households with school-age children associated with ILI-related reactive school closures occurring during eight academic years prior to the COVID-19 pandemic, from 2011−2012 to 2018−2019, in the United States, as well as to explore the costs by academic year, state, and the urbanicity of school location.

## Materials and Methods

### Data sources

We used prospectively collected data on reactive school closures related to ILI from August 1, 2011, to June 30, 2019, for K−12 schools and school districts in the United States (50 states and the D.C.). The data on ILI-related school closures were collected by daily systematic online searches of Google and Lexis-Nexis to identify public announcements of unplanned school closures lasting ≥1 school day [7, 16]. For this analysis, district-wide closure events were disaggregated into their individual schools using data from the National Center for Education Statistics (NCES) (e.g., closure of a school district with 20 schools was represented as 20 closed schools). The duration of school closures was calculated from the date of closure to the date of reopening for each closure event, excluding any planned closures (e.g., weekends or school holidays). The schools with closures were linked with the data from the NCES, which includes the characteristics of each school, such as the total number of students and teachers, grade level, school location, and locale. Details of data collection were summarized in previous studies [7, 16].

### Outcome measures

We estimated the economic cost of ILI-related reactive school closures, including productivity costs for parents, teachers, and non-teaching school staff (hereafter referred to as school staff) associated with the closures. School staff included school district staff (officials and administrators, instruction coordinators, administrative support staff) and school staff excluding teachers (principals and assistant principals, instructional aides, guidance counselors, librarians, school and library support staff, student support staff, and other support services staff) [17]. The average ratio of students to school staff by state and year was used to estimate the lost productivity of school staff during closures [18].

To estimate productivity costs associated with ILI-related school closures, we employed a human-capital approach and considered the hours of productivity lost due to school closures for parents, teachers, and school staff [19]. The productivity cost was estimated by paid work time lost valued at the average wage rate by multiplying the number of days for each closure by the average hourly or daily wage rate. The estimation of productivity costs of parents, teachers, and school staff are explained in the following paragraphs. The summarized estimation equations and the parameter values and assumptions used in the cost estimation are described in Appendices 1-1 and 1-2.

The value of parental work absenteeism was estimated based on survey data from previously examined unplanned school closures related to influenza [11, 13, 20–23]. To estimate the number of parents missing work because of closures, we first estimated the number of households affected by dividing the number of students by the average number of children per household (1.75) [24]. The number of households was then multiplied by the fraction of households with children whose parents missed work; we used the U.S. national-level reported rate of 20% [20] missing work due to influenza for the base case and conducted one-way sensitivity analyses with rates from 10% to 45% [11, 13, 20–23]. Next, the estimated number of parents missing work was multiplied by the state- and year-specific average hourly wage for all occupations (U.S. Bureau of Labor Statistics [BLS] occupation code 00-0000), with an assumption of 8 hours worked per day including non-wage benefits, and by the number of days of closures. The non-wage benefits rate was estimated as a fraction of total wages by year based on the BLS data [25].

For the productivity cost of teachers, we used the number of teachers for each school, which was extracted from the NCES data, and multiplied it by the duration of closure and the state- and year-specific average daily wages of teachers, including non-wage benefits [25]. The average daily wages were estimated by dividing the state- and year-specific average annual wage by 180, the minimum days of instructional time in a school year [26]. The wages for teachers at high schools were based on the average wages for secondary schools (BLS occupation code 25−2031), and the average wages for elementary and middle schools (BLS occupation code 25−2021) were used for teachers of all the other grades.

The NCES dataset did not include the number of school staff for each school. We estimated the number of staff by dividing the number of students by the ratio of students to school staff by state and year [18]. School staff include several occupation categories; we used a weighted average of national daily wage by year by: 1) calculating weights based on the number of school staff for each occupation based on the NCES’s national number of school staff by year [17, 27], 2) multiplying the weight by the reported average yearly wage for each occupation from BLS by year [28], and 3) dividing by 180 schooldays per school year to estimate the daily wage. The NCES categories for school staff and BLS categories for wage estimates included: officials and administrators (BLS occupation code 11-9032), instruction coordinators (BLS occupation code 25-9031), administrative support staff (BLS occupation code 11-9032), principles and assistant principals (average of BLS occupation code 11-9032 and 25-2031), instructional aides (BLS occupation code 25-9031), guidance counselors (BLS occupation code 21-1012), librarians (BLS occupation code 25-4022), school and library support staff (BLS occupation code 25-9099), student support staff (BLS occupation code 21-1021), and other support services staff (BLS occupation code 43-0000). We also considered non-wage benefits and then multiplied it by the number of days of school closures.

### Analysis

We first examined the number of ILI-related reactive school closures by academic year, length of closure, and urbanicity of school location. Descriptive findings are reported for the following measures: (1) total productivity cost and (2) annual total productivity cost per student.

The total productivity cost for each ILI-related school closure was estimated by summing the productivity costs of parents, teachers, and school staff (stratified by academic year from 2011−2012 to 2018−2019 and by state). We also estimated the mean annual total productivity cost and stratified it into two study periods, from 2011−2012 to 2015−2016 and from 2016−2017 to 2018−2019, because the number of closures was much higher in the 2016−2017 to 2018−2019 period than in the 2011−2012 to 2015−2016 period.

The annual total productivity cost per student was estimated at two levels: (1) among public and private schools with ILI-related closures from 2016−2017 to 2018−2019 and (2) among all U.S. public schools from 2011−2012 to 2018−2019. The annual total productivity cost per student among schools with ILI-related closures was estimated for each school by dividing the total productivity costs between 2016−2017 and 2018−2019 by the average number of students per year and the number of academic years with ILI− related closures (e.g., if a school had ILI−related closures in 2016−2017 and 2018−2019, then two years would be used in the denominator. The cost per student among schools with closures were further explored by urbanicity of school location (city, suburban, town, and rural) [29], and we investigated outlier estimates defined as cost greater than 1.5 times the interquartile range above the 75^th^ percentile.

To estimate the annual total productivity cost per student across all U.S. public schools, we excluded the costs of closures for private schools, which accounted for 4% of ILI-related reactive school closures from 2011−2012 to 2018−2019. We then estimated the annual total productivity cost per student for the three time periods (2011−2012 to 2018−2019, 2011−2012 to 2015−2016, and 2016−2017 to 2018−2019), by dividing the total productivity costs by the number of school years (e.g., eight years for the period from 2011−2012 to 2018−2019) and the NCES estimate of the total number of students in U.S. public schools in 2015−2016 by state [30]. We further examined the annual total productivity cost per student by urbanicity of school location and by state for the five highest-cost states during each time period versus the remaining 45 states. The urbanicity of school location was available in NCES datasets [30].

All costs were adjusted to 2019 U.S. dollars using the Consumer Price Index [31]. Analyses were conducted using Microsoft Excel and Stata SE Statistical Software (version 16.1, StataCorp LLC, College Station, TX).

## Results

### ILI-related School Closures

During the eight academic years from 2011−2012 to 2018−2019, there were a total of 5,959 ILI-related school closures among 5,326 schools, with far more in the 2016−2017 to 2018−2019 period (1,711 closures per year) than in the 2011−2012 to 2015−2016 period (165 closures per year) on average (Fig 1 and Appendix 2). Most of the schools with closures experienced one closure per school year (86.8%); a few schools had two (12.8%) or three (0.4%) closures in a given school year (Appendix 3). Most closures lasted less than 4 days (1−day [41%], 2−day [41%], and 3−day [16%]), and the number of closures with ≥4 days was higher during the 2016−2017 to 2018−2019 period (410 closures) than during the 2011−2012 to 2015−2016 period (20 closures) (Appendix 4). Most ILI-related school closures occurred in public schools (96%), among which a majority were located in rural areas (55%) or towns (23%) (Appendix 5-1). Among public schools with closures, most school closures lasting ≥4 days occurred in rural areas (58%) or towns (16%) (Appendix 5-1) and most schools with multiple closures in a given school year also occurred in rural areas (52%) and towns (21%) (Appendix 5-2).

**Fig 1.**
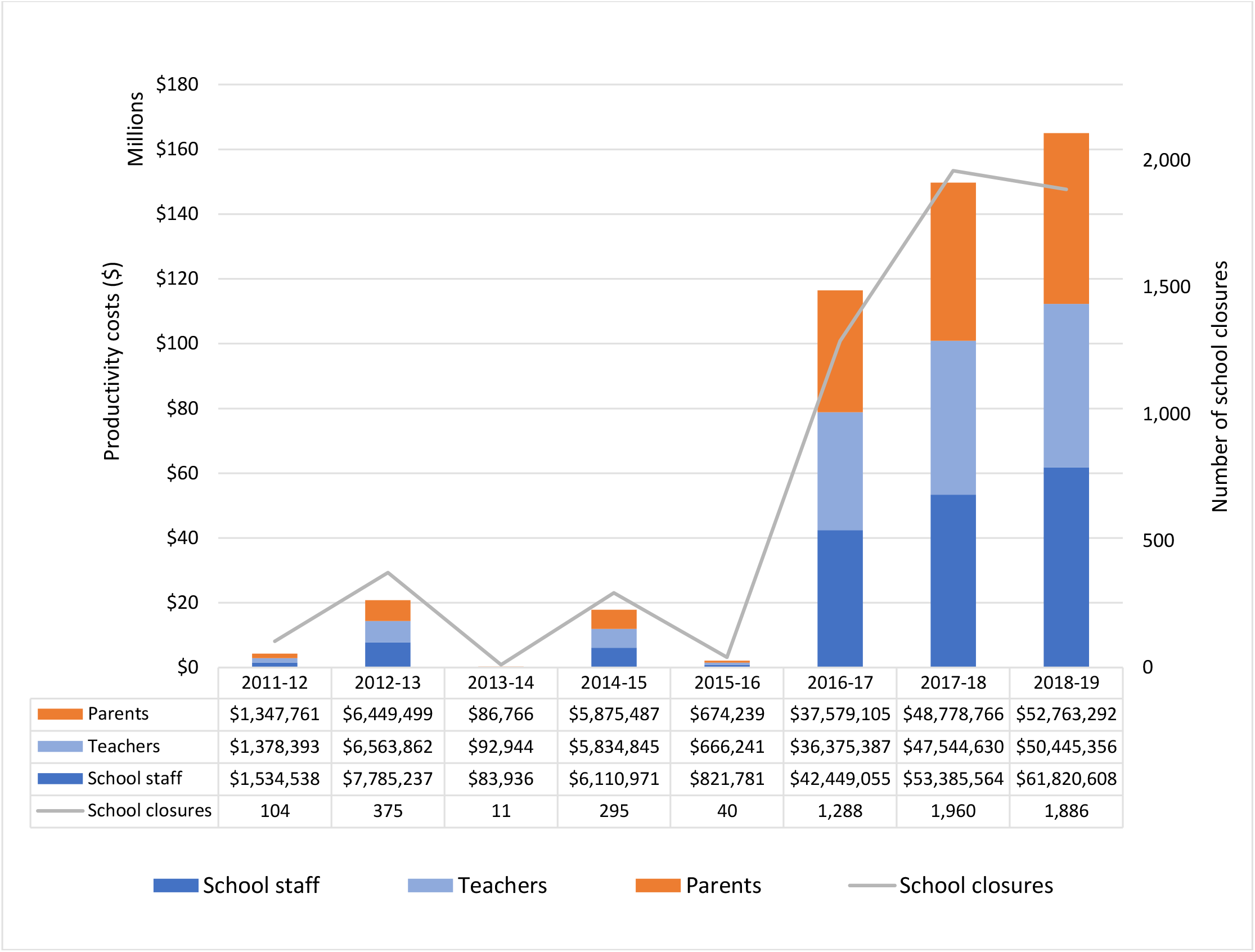
Total productivity costs of reactive school closures associated with ILI by school year from 2011−2012 to 2018−2019 (2019 USD) subdivided by parents, teachers, and school staff. ^a^ Non-teaching school staff category includes school district staff (officials and administrators, instruction coordinators, and administrative support staff) and school staff (principals and assistant principals, instructional aides, guidance counselors, librarians, school and library support staff, student support staff, and other support services staff). (https://nces.ed.gov/programs/digest/d21/tables/dt21_213.20.asp?current=yes) ILI, influenza or influenza-like illness

### Total Productivity Cost

Total productivity costs of school staff, teachers, and parents associated with ILI-related school closures from 2011−2012 to 2018−2019 was estimated at $476 million (Fig 1 and Appendix 6) and ranged from $400 million to $668 million depending on the fraction of parents assumed to miss work due to their children’s school being closed due to ILI (Appendix 7). The total productivity costs were higher in the 2016−2017 to 2018−2019 period than in the 2011−2012 to 2015−2016 period; mean annual costs were $144 million and $9 million, respectively (Appendix 2). The mean productivity costs for parents, teachers, and school staff were $19 million, $18 million, and $22 million, respectively, from 2011−2012 to 2018−2019.

Total productivity costs were highest in states in the South and Midwest, including Tennessee, Kentucky, Texas, Michigan, Oklahoma, Alabama, and Arizona (Fig 2), with most of the costs being incurred in Tennessee (55%) and Kentucky (21%). Among these seven states, the mean annual cost ranged from $1 million in Arizona to $32 million in Tennessee during the 2011−2012 to 2018−2019 period (Appendix 8). These states also had more ILI-related school closures than the other states; the average annual numbers of school closures were 339 in Tennessee, 139 in Kentucky, 47 in Texas, 43 in Michigan, and 39 in Oklahoma. Tennessee and Kentucky had the highest closure costs, with mean costs per year that were 5 to 13 times higher than the third highest state. Additionally, Tennessee and Kentucky accounted for the majority of schools with multiple closures (Appendix 3) and with durations ≥4 days (Appendix 4).

**Fig 2.**
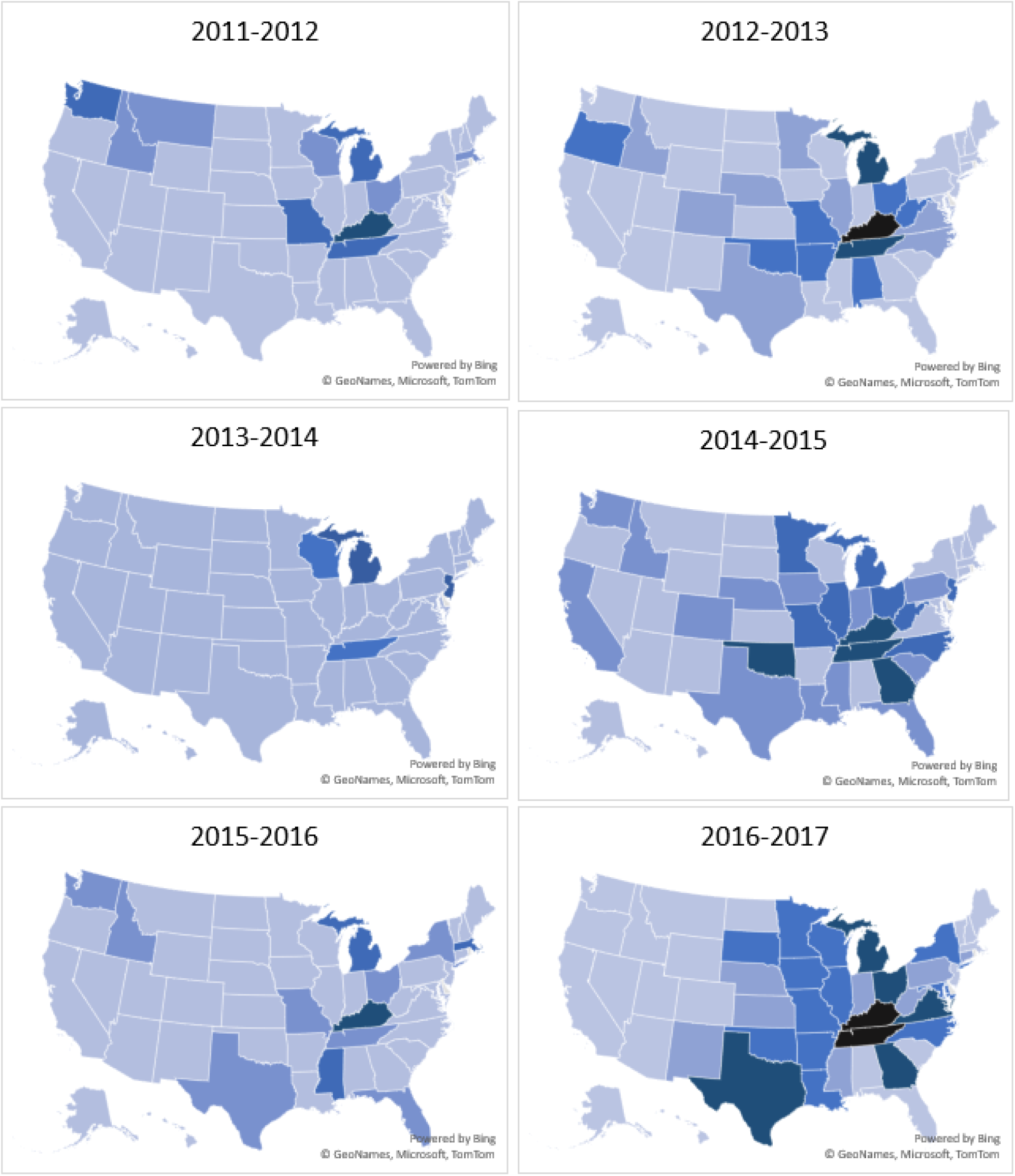
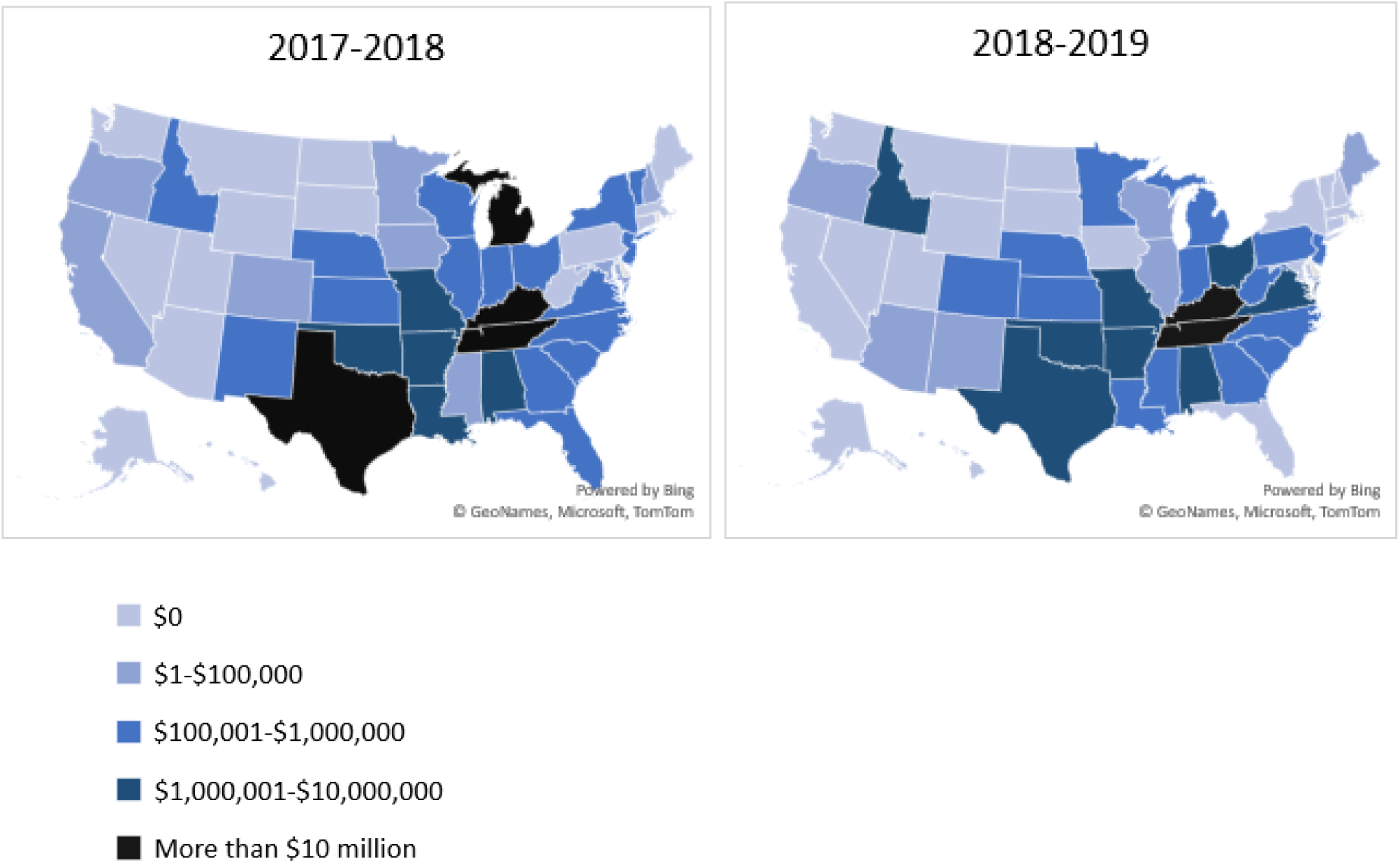
Total productivity costs of reactive school closures associated with ILI by state and school year from 2011−12 to 2018−19 (2019 USD) ILI, influenza or influenza-like illness

Similar trends were observed in state-specific sub-analyses by study period when comparing the 2016− 2017 to 2018−2019 period ($82 million in Tennessee and $28 million in Kentucky) to the 2011−2012 to 2015−2016 period ($2 million and $3 million, respectively).

### Annual Total Productivity Cost Per Student within Schools with ILI-related Closures and Across All Public Schools

Among the schools with ILI-related closures between 2016−2017 and 2018−2019, the annual total productivity cost was estimated at $100 per student (Fig 3 and Appendix 9). When estimating the cost by urbanicity of school location, the schools in the city category had a lower median cost estimate than those in other categories. In addition, there were more outlier estimates among the schools in the rural category (96 schools) than in the others (55, 44, and 42 schools for city, suburban, and town schools, respectively).

**Fig 3.**
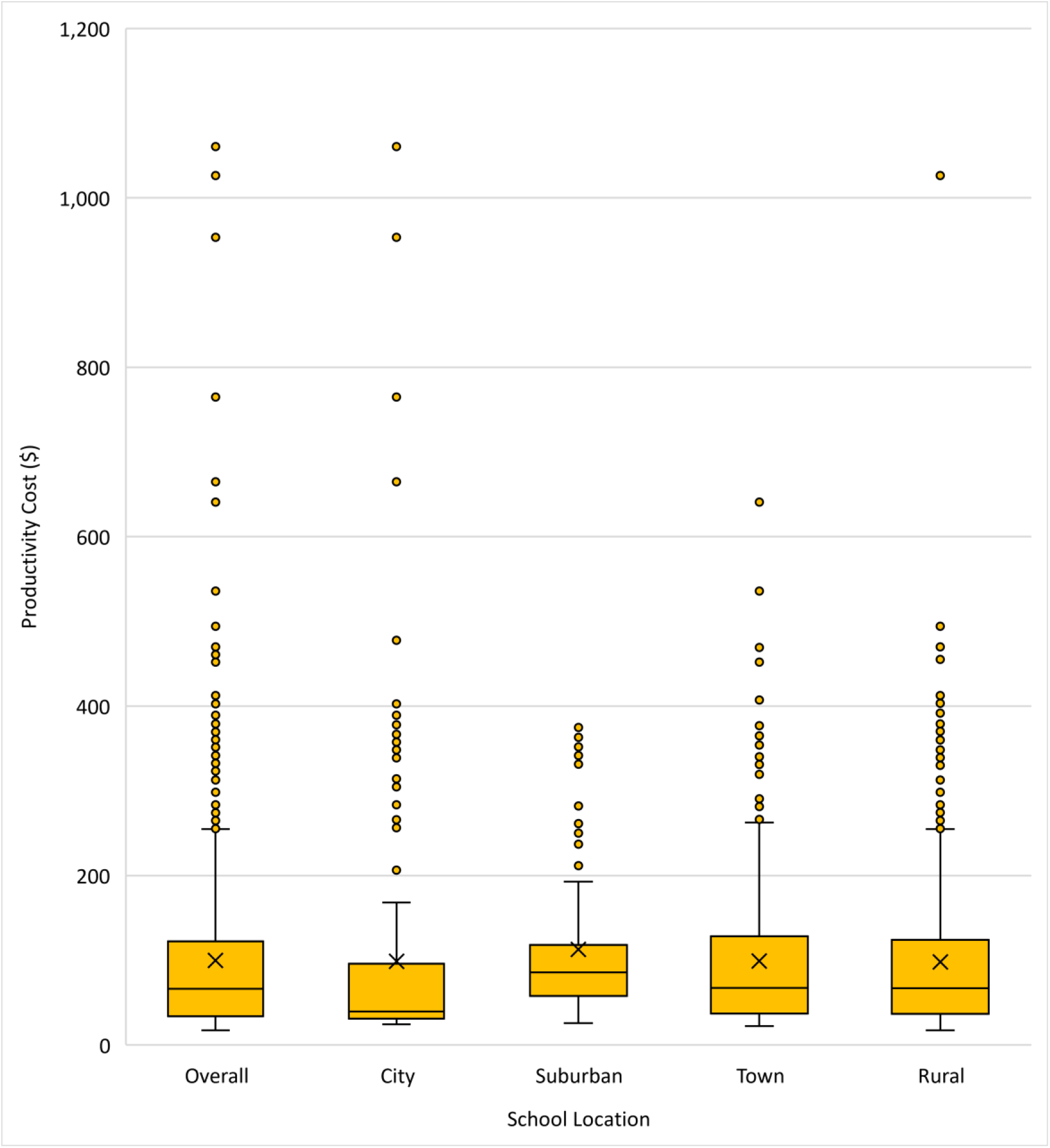
**Annual total productivity cost per student associated with ILI-related reactive school closures among only schools that had closures between the 2016−2017 and 2018−2019, by urbanicity of school location (city, suburban, town, rural, 2019 USD per student-year)** The minimum estimate is shown at the end of the lower whisker line. The 25^th^ percentile is the far bottom of the box. The median (50^th^ percentile) is shown as a line near the center of the box. The mean is shown as X. The 75^th^ percentile is shown as the top of the box. The line extending from the box represents the maximum estimate less than 1.5 times the interquartile range above the 75^th^ percentile, shown as the top of the upper whisker line (i.e., largest observation ≤ 1.5 × (75^th^ percentile − 25^th^ percentile) + 75^th^ percentile). Additional outliers (shown as yellow points) are plotted as point values for schools with estimated costs exceeding a distance of 1.5 times the interquartile range above the 75^th^ percentile. ILI, influenza or influenza-like illness

Among all U.S. public schools (including schools that did not close), the annual total productivity cost was $1.2 per student from 2011−2012 to 2018−2019; the annual cost per student was much higher in Tennessee ($33) and Kentucky ($19) compared to the national average ($1.2) and compared to the other three states in the top five ($2.1 to $2.4 in Arkansas, Oklahoma, and Idaho) (Table 1 and Appendices 10 and 11). Similar trends were observed during both the 2011−2012 to 2015−2016 and 2016−17 to 2018− 2019 periods, with costs in Tennessee and Kentucky that were 14 to 29 times as high as the national average.

**Table 1.** Annual total productivity cost per student associated with ILI-related reactive school closures among U.S. public schools, by urbanicity of school location ^a^ (2019 USD per student-year)

| Academic Year | Overall | By Urbanicity of School Location |  |  |  |
| --- | --- | --- | --- | --- | --- |
|  |  | City | Suburban | Town | Rural |
| **2011–2012 to 2018–2019** |  |  |  |  |  |
| All U.S. | 1.17 | 0.52 | 0.45 | 2.52 | 2.93 |
| Tennessee | 32.50 | 17.25 | 35.36 | 41.20 | 42.27 |
| Kentucky | 18.58 | 7.25 | 3.02 | 21.87 | 30.66 |
| Arkansas | 2.38 | 0.00 | 1.89 | 4.32 | 3.26 |
| Oklahoma | 2.20 | 1.26 | 0.13 | 2.08 | 4.53 |
| Idaho | 2.10 | 0.02 | 0.14 | 2.65 | 5.75 |
| Others <sup>b</sup> | 0.21 | 0.09 | 0.05 | 0.52 | 0.59 |
| **2011–2012 to 2015–2016** |  |  |  |  |  |
| All U.S. | 0.17 | 0.01 | 0.02 | 0.34 | 0.64 |
| Kentucky | 5.13 | 0.00 | 1.09 | 4.98 | 9.91 |
| Tennessee | 2.86 | 0.02 | 0.41 | 4.16 | 6.85 |
| Oklahoma | 0.67 | 0.00 | 0.10 | 0.35 | 1.84 |
| West Virginia | 0.35 | 0.00 | 0.00 | 0.12 | 0.77 |
| Arkansas | 0.33 | 0.00 | 0.12 | 0.00 | 0.93 |
| Others <sup>b</sup> | 0.03 | 0.01 | 0.01 | 0.04 | 0.06 |
| **2016–2017 to 2018–2019** |  |  |  |  |  |
| All U.S. | 2.83 | 1.38 | 1.15 | 6.15 | 6.75 |
| Tennessee | 81.92 | 45.98 | 93.62 | 102.91 | 101.31 |
| Kentucky | 40.99 | 19.33 | 6.22 | 50.04 | 65.25 |
| Arkansas | 5.80 | 0.00 | 4.84 | 11.51 | 7.13 |
| Idaho | 5.39 | 0.06 | 0.15 | 7.08 | 14.75 |
| Alabama | 4.49 | 0.08 | 0.00 | 12.56 | 6.63 |
| Others <sup>b</sup> | 0.50 | 0.26 | 0.12 | 1.11 | 1.41 |

<sup>a</sup> Total number of students in U.S. public schools (overall and by urbanicity category) were based on the National Center for Education Statistics (https://nces.ed.gov/pubs2018/2018052/tables/table_04.asp).
<sup>b</sup> 45 states and the District of Columbia
\$0.00 indicates that there were no reactive school closures related to the ILI.
ILI, influenza or influenza-like illness

When examining the annual total productivity cost per student by urbanicity of school location among all U.S public schools, schools in rural areas ($2.9) or towns ($2.5) had higher costs than those in cities ($0.6) or suburban areas ($0.5). Rural and town areas had higher costs per student relative to city or suburban areas across most state or period subgroup analyses.

## Discussion

While preemptive school closures have been recommended for a severe influenza pandemic as a nonpharmaceutical intervention [3], influenza-related reactive school closures are a consequence of widespread illness in schools [3]; as such, they have occurred both during pandemics [32] and during local seasonal influenza outbreaks [7, 16]. We estimated the economic costs of ILI-related reactive school closures over the eight consecutive inter-pandemic academic years, from 2011−2012 to 2018−2019, in the United States by measuring the productivity costs of parents, teachers, and school staff by state and urbanicity of school location. We found that ILI-related reactive school closures resulted in productivity costs of $476 million in total during the eight academic years and that most of the costs (90%) were incurred in the last three years of the analysis period. The per-student cost burden of public school closures was disproportionately higher in Tennessee and Kentucky than in the other states. In addition, the costs per student were higher for schools in rural areas or towns than in urban or suburban areas across most subgroup analyses by state or period.

The estimated costs associated with reactive ILI-related closures during seasonal influenza outbreaks are substantial (an annual average of $143 million in the 2016−2017 to 2018−2019 period) but significantly lower than the estimated costs of more systematic preemptive school closures that may be considered during influenza pandemics in the United States ($5 to $24 billion for 2-week closures) [33]. This difference is not surprising because the number of schools and the length of closures associated with ILI in this study were significantly fewer (5% of total U.S. schools [34] vs. all U.S. schools) and shorter (average duration of 2 days vs. 2 weeks) than those considered for a hypothetical closure strategy during an influenza pandemic [33]. If we apply our cost per student-day estimates to a scenario in which all U.S. schools would close for two weeks, our cost estimate would be approximately $20 billion, which is in line with the previous estimates for two-week preemptive closures during an influenza pandemic [33]. However, a major difference should be noted here: while appropriately timed and targeted preemptive school closures could reduce influenza transmission in surrounding communities in a cost-effective manner [35], reactive closures occur after substantively increased student absenteeism, i.e., too late to have a desirable epidemiologic impact while still carrying a substantial cost for families and society. In addition, the costs of school closures may be reduced after the COVID-19 pandemic due to increased availability of distancing learning; approximately 93% of households with school-age children had participated in distance learning by September 2020 during school closures due to COVID-19 [36].

The cost burden of ILI-related reactive school closures was unevenly distributed with regard to academic year and geography. Specifically, higher costs of ILI-related closures in the 2016−2017 to 2018−2019 period (vs. the 2011−2012 to 2015−2016 period) and in Tennessee and Kentucky (vs. the other states) were primarily due to the greater percentage of affected schools and longer duration of closures. The temporal patterns reflect the year-to-year variation in seasonal influenza outbreaks; it has been reported that a higher number and longer duration of closures were associated with the seasons dominated by influenza A virus subtype H3N2 [37]. Additionally, 2017−2018 has been reported as an H3N2−predominant influenza season with high severity, record hospitalization rates, and a high number of influenza-associated pediatric deaths [38]. Efforts to improve influenza active monitoring and surveillance by health authorities and improving influenza vaccine coverage may reduce the costs of reactive school closures. As the effectiveness of seasonal influenza vaccines is typically found to be lower against the influenza A virus subtype H3N2, it would be useful to develop more broadly protective and longer-lasting influenza vaccines by increasing antigen content or adding adjuvants [39].

ILI-related reactive school closures disproportionately affected public school students in rural areas and towns. The number of ILI-related school closures in these areas was four times as high as those in cities or suburban areas, with more of these schools having multiple closures or longer duration closures, resulting in a cost per student estimate that was five-times greater in rural areas or towns compared to cities or suburban areas. The higher frequency of ILI-related school closures could be partially related to lower influenza vaccination rates among children residing in rural areas compared to cities or suburban areas during the 2011−2012 through 2018−2019 influenza seasons [40]. In addition, given the high level of income inequality in rural areas [41], our findings suggest that the ILI-related school closures may have added to the economic burden on rural households, especially low-income ones. This may be in line with a previous finding that reduced income from missed work was the most frequently reported reason for economic difficulties during ILI-related school closures [11]. Furthermore, students missing meals was reported as another difficulty [11]; as public schools provide eligible students with free or reduced-price lunches [42], ILI-related school closures could cause food insecurity problems for low-income students. This could be a bigger problem in rural areas or towns where a relatively large number of low-income students live [41]. Therefore, additional efforts may be needed to reduce rural-urban disparities in ILI-related school closures and their associated cost burden by understanding factors contributing to the potential disparities in influenza incidence and childhood vaccination coverage in rural areas, as well as by developing strategies to reduce the financial burden of school closures.

This study is subject to several limitations. First, the data on ILI-related school closures may not be complete or entirely accurate due to the following reasons: 1) it was limited to school closures found in publicly available online sources and some closures might have been missed, particularly those announced solely through non-public methods (email, text message, etc.); 2) it included 50 U.S. states and D.C. but excluded U.S. territories, and 3) it was limited to the school closures announced in English. Second, due to a limited number of available studies, the proportion of parents who missed work due to influenza used in this study was based on a single study conducted during the 2009 influenza A (H1N1) pandemic and may not be representative of locations where reactive closures for seasonal influenza outbreaks occur. This study also may not account for regional variation in parents’ work absenteeism by state or urbanicity. However, since we used an estimate from a national sample of parents who experienced school dismissals related to influenza pandemic [20], our total cost estimates should be relatively reliable as a median estimate (see Appendix 7 for lower and upper bound estimates of productivity costs). Third, our cost estimates may underestimate the cost burden associated with ILI-related school closures regarding the extent of caregivers’ efforts to make alternative childcare arrangements or students losing access to subsidized school lunches. We did not attempt to account for non-productivity costs to households including payments to childcare providers during reactive school closures. Fourth, because we estimated wages for teachers and school staff using the average wage by occupation from BLS statistics and the overall average wage across all occupations for parents; our cost estimates do not account for differences in wage rates by urbanicity or may not account for differences in socioeconomic differences between schools that close versus those that remain open during local influenza outbreaks.

## Conclusions

To the best of our knowledge, this is the first study that has estimated the economic costs associated with reactive ILI-related closures. We found that ILI-related school closures occurring over the eight academic years prior to the COVID-19 pandemic (2011−2012 to 2018−2019) caused a substantial economic cost burden, which was particularly pronounced in 2016−2017, 2017−2018, and 2018−2019. Additionally, the school closures exerted a disproportionate economic burden on families in rural areas or towns and especially in the states of Tennessee and Kentucky where a higher percentage of schools closed, and many were closed for four days or more. As school closures vary with the severity of seasonal influenza, holistic strategies may be necessary to sustainably reduce the seasonal influenza attack rates in schools and minimize the need for and cost of ILI-related reactive school closures which, because of their late timing relative to already advanced spread of influenza in school, have little effect on further transmission [11, 32]. A better understanding of the reasons for the higher occurrence of and high cost of ILI-related closures among students in rural areas or towns, especially in the states with exceptionally high burden (Tennessee and Kentucky), is needed to reduce the gaps in health and economic disparities between rural/town and city/suburban areas.

## Data Availability

All relevant data are available from the Data.CDC.gov database (https://data.cdc.gov/Public-Health-Surveillance/Prolonged-Unplanned-School-Closures-USA-2011-2019/5iuf-feyd).

## Supporting information

**Appendix 1-1**. Equations for estimating productivity costs of parents, teachers, and non-teaching school staff

**Appendix 1-2**. Parameters values and assumptions for productivity costs estimation

**Appendix 2**. Mean annual number of ILI-related reactive school closures and mean annual productivity costs, by study period

**Appendix 3**. Number of schools with multiple ILI-related reactive school closures, from 2011-2012 to 2018-2019

**Appendix 4**. Number of ILI-related reactive school closures by length of closures and academic year

**Appendix 5-1**. Number of ILI-related reactive school closures among public schools from 2011-2012 to 2018-2019, by urbanicity of school location and length of closures

**Appendix 5-2**. Number of public schools with multiple ILI-related reactive school closures, by urbanicity of school location

**Appendix 6**. Number of ILI-related reactive school closures and productivity costs from 2011-2012 to 2018-2019

**Appendix 7**. Sensitivity analysis of productivity costs of parents and total productivity costs associated with ILI-related reactive school closures by varying the fraction of parents missing work (range: 10% to 45%)

**Appendix 8**. Mean number of ILI-related reactive school closures per year and the mean total productivity cost per year, by state and by study period

**Appendix 9**. Annual total productivity cost per student among schools with ILI-related reactive closures from 2016−2017 to 2018−2019 (2019 USD)

**Appendix 10**. Total productivity costs associated with ILI-related reactive school closures from 2011− 2012 to 2018−2019 among public schools and total number of students in U.S. public schools in 2015− 2016

**Appendix 11**. Annual total productivity cost per student associated with ILI-related reactive school closures among all U.S. public schools, by state* and academic year (2019 USD)

