## Appendices for "Productivity costs associated with reactive school closures related to influenza or influenza-like illness in the United States from 2011 to 2019"

**Appendix 1-1.** Equations for estimating productivity costs of parents, teachers, and non-teaching school staff

$$Productivity Cost of Parents=\left[ \left( number of students\div average number of children per household \right) \right] \times fraction of households with children whose parents missed work \times average hourly wages of parents by state and year\times8 hours\times\left( 1+ nonwage benefits rate \right)\times number of days of unplanned school closure$$

$$Productivity Cost of Teachers =number of teachers \times average daily wages of teachers by school grade, state, and year \times\left( 1+nonwage benefits rate \right)\times number of days of school closure$$

$$Productivity Cost of Non-teaching School Staff =number of students\div ratio of students to school staff by state and year \times average daily wages of school staff by school grade, state, and year \times\left( 1+nonwage benefits rate \right)\times number of days of school closure$$

**Appendix 1-2.** Parameters values and assumptions for productivity costs estimation

| **Parameter Description** | **Base Value** | **Range** | **Source** |
| --- | --- | --- | --- |
| State- and year-specific average hourly wages for parents  (2019 USD; BLS code 00-0000) | - 2012: $18.9‒$40.7 - 2013: $19.0‒$40.6 - 2014: $19.2‒$40.8 - 2015: $19.5‒$41.6 - 2016: $19.6‒$42.5 - 2017: $19.5‒$43.0 - 2018: $19.3‒$43.0 - 2019: $19.3‒$43.2   (varies by state) | NA | [1] |
| State- and year-specific average annual wages for elementary and middle school teachers (2019 USD; BLS code 25-2021) | - 2012: $44,429‒$82,744 - 2013: $44,335‒$80,154 - 2014: $44,287‒$80,809 - 2015: $45,042‒$81,650 - 2016: $43,833‒$82,372 - 2017: $42,273‒$84,003 - 2018: $41,182‒$84,512 - 2019: $44,060‒$82,830   (varies by state) | NA | [1] |
| State- and year-specific average annual wages for high school teachers (2019 USD; BLS code 25-2031) | - 2012: $44,618‒$82,544 - 2013: $44,478‒$82,579 - 2014: $45,853‒$82,807 - 2015: $46,337‒$86,881 - 2016: $45,228‒$87,368 - 2017: $43,681‒$89,093 - 2018: $43,310‒$86,844 - 2019: $44,610‒$87,240   (varies by state) | NA | [1] |
| Weighted average of national daily wage for non-teaching school staff (2019 USD, varies by year) | $56,646‒$59,495 | NA | [1-3] |
| Staff-student ratio | 1:8 | NA | [4] |
| Average children per household | 1.75 | NA | [5] |
| Fraction of households with children who missed work during unplanned school closures | 20% [6] | 10%–45% | [6-11] |
| Nonwage benefits as a fraction of total wage + nonwage pay for employees (varies by year) | 44.5%‒46.4% | NA | [12] |

USD: US dollars, BLS: U.S. Bureau of Labor Statistics, NA: Not applicable

**Appendix 2.** Mean annual number of ILI-related reactive school closures and mean annual productivity costs, by study period

|  | **2011/12‒2015/16** | **2016/17‒2018/19** | **2011/12‒2018/19** |
| --- | --- | --- | --- |
| **Mean annual number of school closure (n)** | 165 | 1,711 | 745 |
| **Mean annual productivity cost (2019 USD)** |  |  |  |
| Overall | 9,061,300 | 143,713,921 | 59,556,033 |
| School staff | 3,267,293 | 52,551,742 | 21,748,961 |
| Teachers | 2,907,257 | 44,788,458 | 18,612,707 |
| Parents | 2,886,751 | 46,373,721 | 19,194,364 |

ILI, influenza or influenza-like illness

**Appendix 3.** Number of schools with multiple ILI-related reactive school closures, from 2011-2012 to 2018-2019

|  | n | % |
| --- | --- | --- |
| Number of school closures in a given school year | | |
| 1 | 3,038 | 86.8 |
| 2 | 448^a^ | 12.8 |
| 3 | 15^b^ | 0.4 |
| Total | 3,501 | 100 |
| Number of school closures from 2011-12 to 2018-19 | | |
| 1 | 2,184 | 62.4 |
| 2 | 689^c^ | 19.7 |
| 3 | 357^d^ | 10.2 |
| 4 | 94^e^ | 2.7 |
| 5 | 155^f^ | 4.4 |
| 7 | 3^f^ | 0.1 |
| 8 | 17^f^ | 0.5 |
| 9 | 2^f^ | 0.1 |
| Total | 3,501 | 100 |

^a^ Among 448 schools, 274 schools (61%) were located in Tennessee, 83 schools (19%) were in Kentucky, 19 schools (4%) in Texas, 18 schools (4%) in Michigan, and 13 schools (3%) were in Alabama.

^b^ Among 15 schools, 10 schools (67%) were located in Kentucky, 5 schools (33%) were in Tennessee.

^c^ Among 689 schools, 381 schools (55%) were located in Tennessee, 119 schools (17%) were in Kentucky, 38 schools (6%) in Texas, 37 schools (5%) in Michigan, and 30 schools (4%) were in Oklahoma.

^d^ Among 357 schools, 202 schools (57%) were located in Tennessee and 114 schools (32%) were in Kentucky.

^e^ Among 94 schools, 44 schools (47%) were located in Kentucky and 43 schools (46%) were in Tennessee.

^f^ All schools (100%) are located in Tennessee.

ILI, influenza or influenza-like illness

**Appendix 4.** Number of ILI-related reactive school closures by length of closures and academic year

| Academic Year |  | Length of school closures (per closure) ^a^ | | | | | | | |
| --- | --- | --- | --- | --- | --- | --- | --- | --- | --- |
|  | n or % | 1-day | 2-day | 3-day | 4-day | 5-day | 6-day | 7-day | Total |
| 2011-2012 | n | 81 | 23 | 0 | 0 | 0 | 0 | 0 | 104 |
|  | % | 77.9 | 22.1 | 0.0 | 0.0 | 0.0 | 0.0 | 0.0 | 100 |
| 2012-2013 | n | 218 | 104 | 33 | 20 | 0 | 0 | 0 | 375 |
|  | % | 58.1 | 27.7 | 8.8 | 5.3 | 0.0 | 0.0 | 0.0 | 100 |
| 2013-2014 | n | 11 | 0 | 0 | 0 | 0 | 0 | 0 | 11 |
|  | % | 100.0 | 0.0 | 0.0 | 0.0 | 0.0 | 0.0 | 0.0 | 100 |
| 2014-2015 | n | 157 | 102 | 36 | 0 | 0 | 0 | 0 | 295 |
|  | % | 53.2 | 34.6 | 12.2 | 0.0 | 0.0 | 0.0 | 0.0 | 100 |
| 2015-2016 | n | 24 | 12 | 4 | 0 | 0 | 0 | 0 | 40 |
|  | % | 60.0 | 30.0 | 10.0 | 0.0 | 0.0 | 0.0 | 0.0 | 100 |
| 2016-2017 | n | 516 | 462 | 155 | 129 | 17 | 9 | 0 | 1,288 |
|  | % | 40.1 | 35.9 | 12.0 | 10.0 | 1.3 | 0.7 | 0.0 | 100 |
| 2017-2018 | n | 810 | 883 | 162 | 47 | 29 | 27 | 2 | 1,960 |
|  | % | 41.3 | 45.1 | 8.3 | 2.4 | 1.5 | 1.4 | 0.1 | 100 |
| 2018-2019 | n | 594 | 848 | 294 | 93 | 35 | 22 | 0 | 1,886 |
|  | % | 31.5 | 45.0 | 15.6 | 4.9 | 1.9 | 1.2 | 0.0 | 100 |
| Total | n | 2,411 | 2,434 | 684 | 289^b^ | 81^b^ | 58^b^ | 2^b^ | 5,959^c^ |
|  | % | 40.5 | 40.9 | 11.5 | 4.9 | 1.4 | 1.0 | 0.0 | 100 |

^a^ Closures with missing information on the length of closures were imputed to 1-day (n=631, 11% of total closures).

^b^ Among a total of 430 closures with ≥4 days, 195 (45%) occurred in Tennessee, 156 (36%) in Kentucky, 16 (4%) in Alabama, 12 (3%) in Missouri, and 11 (3%) in Texas.

^c^ During the eight academic years from 2011-12 to 2018-19, 5,959 ILI-related closures occurred among 3,501 schools.

ILI, influenza or influenza-like illness

**Appendix 5-1.** Number of ILI-related reactive school closures among public schools from 2011-2012 to 2018-2019, by urbanicity of school location and length of closures

|  | City | | Suburban | | Town | | Rural | | Total | |
| --- | --- | --- | --- | --- | --- | --- | --- | --- | --- | --- |
|  | n | % | n | % | n | % | n | % | n | % |
| By Length of closures (per closure) | | | | | | | | | | |
| 1-day | 422 | 55.1 | 225 | 38.0 | 429 | 33.6 | 1,214 | 39.3 | 2,290 | 40.0 |
| 2-day | 269 | 35.1 | 282 | 47.6 | 572 | 44.8 | 1,234 | 40.0 | 2,357 | 41.2 |
| 3-day | 15 | 2.0 | 37 | 6.3 | 212 | 16.6 | 401 | 13.0 | 665 | 11.6 |
| 4-day | 51 | 6.7 | 48 | 8.1 | 28 | 2.2 | 147 | 4.8 | 274 | 4.8 |
| 5-day | 9 | 1.2 | 0 | 0.0 | 17 | 1.3 | 53 | 1.7 | 79 | 1.4 |
| 6-day | 0 | 0.0 | 0 | 0.0 | 20 | 1.6 | 37 | 1.2 | 57 | 1.0 |
| 7-day | 0 | 0.0 | 0 | 0.0 | 0 | 0.0 | 2 | 0.1 | 2 | 0.0 |
| Total | 766 | 100.0 | 592 | 100.0 | 1,278 | 100.0 | 3,088 | 100.0 | 5,724* | 100.0 |

* During the eight academic years from 2011-12 to 2018-19, 5,724 ILI-related closures (96% of total closures) occurred among 3,289 public schools.

ILI, influenza or influenza-like illness

**Appendix 5-2.** Number of public schools with multiple ILI-related reactive school closures, by urbanicity of school location

|  | City | | Suburban | | Town | | Rural | | Total | |
| --- | --- | --- | --- | --- | --- | --- | --- | --- | --- | --- |
|  | n | % | n | % | n | % | n | % | n | % |
| Number of closures for each school in a given school year | | | | | | | | | | |
| 1 | 357 | 85.6 | 224 | 78.9 | 673 | 87.4 | 1,579 | 86.9 | 2,833 | 86.1 |
| 2 | 60 | 14.4 | 60 | 21.1 | 88 | 11.4 | 233 | 12.8 | 441 | 13.4 |
| 3 | 0 | 0.0 | 0 | 0.0 | 9 | 1.2 | 6 | 0.3 | 15 | 0.5 |
| Total | 417 | 100.0 | 284 | 100.0 | 770 | 100.0 | 1,818 | 100.0 | 3,289 | 100.0 |
| Number of closures for each school from 2011-2012 to 2018-2019 | | | | | | | | | | |
| 1 | 263 | 63.1 | 133 | 46.8 | 466 | 60.5 | 1,127 | 62.0 | 1,989 | 60.5 |
| 2 | 55 | 13.2 | 67 | 23.6 | 196 | 25.5 | 359 | 19.7 | 677 | 20.6 |
| 3 | 51 | 12.2 | 43 | 15.1 | 60 | 7.8 | 199 | 10.9 | 353 | 10.7 |
| 4 | 0 | 0.0 | 12 | 4.2 | 23 | 3.0 | 58 | 3.2 | 93 | 2.8 |
| 5 | 48 | 11.5 | 29 | 10.2 | 23 | 3.0 | 55 | 3.0 | 155 | 4.7 |
| 7 | 0 | 0.0 | 0 | 0.0 | 0 | 0.0 | 3 | 0.2 | 3 | 0.1 |
| 8 | 0 | 0.0 | 0 | 0.0 | 2 | 0.3 | 15 | 0.8 | 17 | 0.5 |
| 9 | 0 | 0.0 | 0 | 0.0 | 0 | 0.0 | 2 | 0.1 | 2 | 0.1 |
| Total | 417 | 100.0 | 284 | 100.0 | 770 | 100.0 | 1,818 | 100.0 | 3,289 | 100.0 |

ILI, influenza or influenza-like illness

**Appendix 6.** Number of ILI-related reactive school closures and productivity costs from 2011-2012 to 2018-2019

|  | Number of school closures | Productivity Costs (2019 USD) | | | |
| --- | --- | --- | --- | --- | --- |
| School year |  | Parents | Teachers | School staff | Total |
| 2011-2012 | 104 | 1,347,761 | 1,378,393 | 1,534,538 | 4,260,692 |
| 2012-2013 | 375 | 6,449,499 | 6,563,862 | 7,785,237 | 20,798,599 |
| 2013-2014 | 11 | 86,766 | 92,944 | 83,936 | 263,646 |
| 2014-2015 | 295 | 5,875,487 | 5,834,845 | 6,110,971 | 17,821,303 |
| 2015-2016 | 40 | 674,239 | 666,241 | 821,781 | 2,162,261 |
| 2016-2017 | 1,288 | 37,579,105 | 36,375,387 | 42,449,055 | 116,403,546 |
| 2017-2018 | 1,960 | 48,778,766 | 47,544,630 | 53,385,564 | 149,708,960 |
| 2018-2019 | 1,886 | 52,763,292 | 50,445,356 | 61,820,608 | 165,029,256 |
| Total | 5,959 | 153,554,916 | 148,901,658 | 173,991,690 | 476,448,263 |

ILI, influenza or influenza-like illness

**Appendix 7.** Sensitivity analysis of productivity costs of parents and total productivity costs associated with ILI-related reactive school closures by varying the fraction of parents missing work (range: 10% to 45%)

|  | Productivity cost of parents (2019 USD) | | | Total productivity costs* (2019 USD) | | |
| --- | --- | --- | --- | --- | --- | --- |
| School year | Base case (20%) | LL (10%) | UL (45%) | Base case (20%) | LL (10%) | UL (45%) |
| 2011‒2012 | 1,347,761 | 673,880 | 3,032,462 | 4,260,692 | 3,586,812 | 5,945,393 |
| 2012‒2013 | 6,449,499 | 3,224,750 | 14,511,373 | 20,798,599 | 17,573,849 | 28,860,473 |
| 2013‒2014 | 86,766 | 43,383 | 195,224 | 263,646 | 220,263 | 372,104 |
| 2014‒2015 | 5,875,487 | 2,937,744 | 13,219,847 | 17,821,303 | 14,883,559 | 25,165,662 |
| 2015‒2016 | 674,239 | 337,120 | 1,517,038 | 2,162,261 | 1,825,141 | 3,005,059 |
| 2016‒2017 | 37,579,105 | 18,789,553 | 84,552,986 | 116,403,546 | 97,613,994 | 163,377,428 |
| 2017‒2018 | 48,778,766 | 24,389,383 | 109,752,223 | 149,708,960 | 125,319,577 | 210,682,417 |
| 2018‒2019 | 52,763,292 | 26,381,646 | 118,717,407 | 165,029,256 | 138,647,609 | 230,983,371 |
| Total | 153,554,916 | 76,777,458 | 345,498,560 | 476,448,263 | 399,670,805 | 668,391,907 |

* Total productivity costs include productivity costs of parents, teachers, and non-teaching school staff.

ILI, influenza or influenza-like illness; LL, lower-level; UL, upper-level

**Appendix 8.** Mean number of ILI-related reactive school closures per year and the mean total productivity cost per year, by state and by study period

|  | **Average number of closures per year (n)** | | | **Average total productivity cost per year (2019 USD)** | | |
| --- | --- | --- | --- | --- | --- | --- |
| **State** | **2011/12‒2015/16** | **2016/17‒2018/19** | **2011/12‒2018/19** | **2011/12‒2015/16** | **2016/17‒2018/19** | **2011/12‒2018/19** |
| TN | 51.0 | 820.0 | 339.4 | 2,947,167 | 82,609,824 | 32,820,664 |
| KY | 52.0 | 282.7 | 138.5 | 3,468,643 | 27,944,236 | 12,646,990 |
| TX | 0.6 | 123.3 | 46.6 | 25,358 | 6,502,554 | 2,454,307 |
| MI | 11.2 | 95.7 | 42.9 | 419,307 | 5,075,112 | 2,165,234 |
| OK | 15.0 | 77.7 | 38.5 | 462,395 | 3,324,601 | 1,535,722 |
| AL | 0.2 | 47.3 | 17.9 | 26,155 | 3,685,264 | 1,398,321 |
| AR | 2.8 | 50.0 | 20.5 | 161,684 | 2,850,305 | 1,169,917 |
| GA | 3.8 | 11.7 | 6.8 | 478,402 | 1,761,701 | 959,639 |
| MO | 7.2 | 38.0 | 2.6 | 261,630 | 1,805,814 | 840,699 |
| ID | 1.8 | 32.3 | 13.3 | 35,110 | 1,548,790 | 602,740 |
| OH | 4.2 | 14.3 | 8.0 | 165,256 | 1,063,443 | 502,076 |
| VA | 0.2 | 15.7 | 6.0 | 1,435 | 1,112,024 | 417,906 |
| LA | 0.2 | 17.0 | 6.5 | 7,348 | 733,123 | 279,514 |
| NC | 3.2 | 12.7 | 6.8 | 90,152 | 510,638 | 247,834 |
| IL | 0.8 | 7.7 | 3.4 | 62,002 | 395,937 | 187,227 |
| MS | 0.4 | 6.3 | 2.6 | 36,370 | 321,805 | 143,408 |
| KS | NR | 11.3 | 4.3 | 0 | 348,436 | 130,664 |
| NJ | 0.8 | 2.0 | 1.3 | 55,570 | 246,436 | 127,145 |
| WI | 0.6 | 7.0 | 3.0 | 8,767 | 278,577 | 109,946 |
| WV | 2.2 | 3.0 | 2.5 | 97,232 | 127,939 | 108,747 |
| CO | 1.0 | 4.3 | 2.3 | 12,838 | 226,404 | 92,925 |
| MN | 1.2 | 2.0 | 1.5 | 38,187 | 141,137 | 76,793 |
| IN | 0.2 | 3.0 | 1.3 | 11,338 | 148,329 | 62,710 |
| NM | NR | 6.7 | 2.5 | 0 | 166,287 | 62,358 |
| NE | 0.6 | 3.7 | 1.8 | 24,522 | 102,175 | 53,642 |
| SC | 0.2 | 2.0 | 0.9 | 14,386 | 97,568 | 45,579 |
| NY | 0.2 | 2.3 | 1.0 | 3,832 | 112,110 | 44,436 |
| FL | 0.4 | 1.7 | 0.9 | 14,516 | 72,301 | 36,185 |
| WA | 1.0 | NR | 0.6 | 56,705 | 0 | 35,441 |
| MD | NR | 1.0 | 0.4 | 0 | 84,814 | 31,805 |
| PA | 0.2 | 0.7 | 0.4 | 4,738 | 70,533 | 29,411 |
| IA | 0.2 | 1.7 | 0.8 | 10,369 | 58,798 | 28,529 |
| OR | 0.2 | 0.7 | 0.4 | 20,078 | 42,588 | 28,520 |
| VT | NR | 1.3 | 0.5 | 0 | 52,341 | 19,628 |
| MA | 0.4 | NR | 0.3 | 28,374 | 0 | 17,734 |
| SD | NR | 3.0 | 1.1 | 0 | 34,879 | 13,080 |
| CA | 0.2 | 0.3 | 0.3 | 2,136 | 22,824 | 9,894 |
| NH | NR | 0.3 | 0.1 | 0 | 13,139 | 4,927 |
| ME | NR | 0.3 | 0.1 | 0 | 8,459 | 3,172 |
| CT | NR | 0.3 | 0.1 | 0 | 7,142 | 2,678 |
| AZ | NR | 0.3 | 0.1 | 0 | 5,534 | 2,075 |
| MT | 0.6 | NR | 0.4 | 3,150 | 0 | 1,969 |
| AK | NR | NR | NR | 0 | 0 | 0 |
| DC | NR | NR | NR | 0 | 0 | 0 |
| DE | NR | NR | NR | 0 | 0 | 0 |
| HI | NR | NR | NR | 0 | 0 | 0 |
| ND | NR | NR | NR | 0 | 0 | 0 |
| NV | NR | NR | NR | 0 | 0 | 0 |
| RI | NR | NR | NR | 0 | 0 | 0 |
| UT | NR | NR | NR | 0 | 0 | 0 |
| WY | NR | NR | NR | 0 | 0 | 0 |

ILI, influenza or influenza-like illness; NR, not reported

**Appendix 9.** Annual total productivity cost per student among schools with ILI-related reactive closures from 2016‒2017 to 2018‒2019 (2019 USD)

|  | **Overall** | **By urbanicity of school location** | | | |
| --- | --- | --- | --- | --- | --- |
| **Estimates** |  | **City** | **Suburban** | **Town** | **Rural** |
| Minimum | 17.3 | 24.3 | 25.9 | 22.2 | 17.3 |
| 25th percentile | 34.0 | 31.3 | 57.8 | 37.2 | 36.7 |
| Median | 66.3 | 39.4 | 85.8 | 67.5 | 66.9 |
| **Mean** | **99.8** | **98.6** | **112.7** | **99.2** | **98.1** |
| 75th percentile | 122.4 | 95.6 | 118.3 | 128.4 | 124.0 |
| Interquartile range | 88.4 | 64.4 | 60.5 | 91.2 | 87.3 |

ILI, influenza or influenza-like illness

**Appendix 10**. Total productivity costs associated with ILI-related reactive school closures from 2011‒2012 to 2018‒2019 among public schools and total number of students in U.S. public schools in 2015‒2016

|  | Overall | By urbanicity of school location | | | |
| --- | --- | --- | --- | --- | --- |
|  |  | City | Suburban | Town | Rural |
| Total productivity costs among public schools with ILI-related closures, 2011-2012 to 2018-2019 (2019 USD) | | | | | |
| All public schools | 460,599,747 | 62,149,629 | 69,812,689 | 112,376,093 | 216,261,335 |
| Tennessee | 257,862,754 | 44,485,510 | 57,512,449 | 54,250,622 | 101,614,173 |
| Kentucky | 100,574,776 | 6,279,838 | 3,608,591 | 30,200,798 | 60,590,412 |
| Arkansas | 9,359,337 | 0 | 1,053,703 | 4,004,342 | 4,301,292 |
| Oklahoma | 12,191,411 | 1,661,003 | 154,842 | 2,718,244 | 7,657,322 |
| Idaho | 4,810,672 | 11,248 | 90,824 | 1,453,958 | 3,265,889 |
| Others (46 states) | 75,800,797 | 9,712,031 | 7,392,280 | 19,748,128 | 38,832,248 |
| Number of students in the U.S. public schools, 2015-2016 (n)* | | | | | |
| All U.S. public schools | 49,312,454 | 14,892,361 | 19,577,044 | 5,572,307 | 9,221,429 |
| Tennessee | 991,648 | 322,286 | 203,288 | 164,614 | 300,469 |
| Kentucky | 676,793 | 108,287 | 149,571 | 172,582 | 247,029 |
| Arkansas | 491,390 | 140,538 | 69,777 | 115,968 | 165,107 |
| Oklahoma | 692,546 | 164,133 | 153,745 | 163,441 | 211,227 |
| Idaho | 286,447 | 66,742 | 79,632 | 68,461 | 71,039 |
| Others (46 states) | 46,173,630 | 14,090,376 | 18,921,030 | 4,887,242 | 8,226,558 |

* Total number of students in the U.S. public schools (overall and by urbanicity category) were based on the National Center for Education Statistics (https://nces.ed.gov/pubs2018/2018052/tables/table_04.asp)

ILI, influenza or influenza-like illness

**Appendix 11**. Annual total productivity cost per student associated with ILI-related reactive school closures among all U.S. public schools, by state* and academic year (2019 USD)

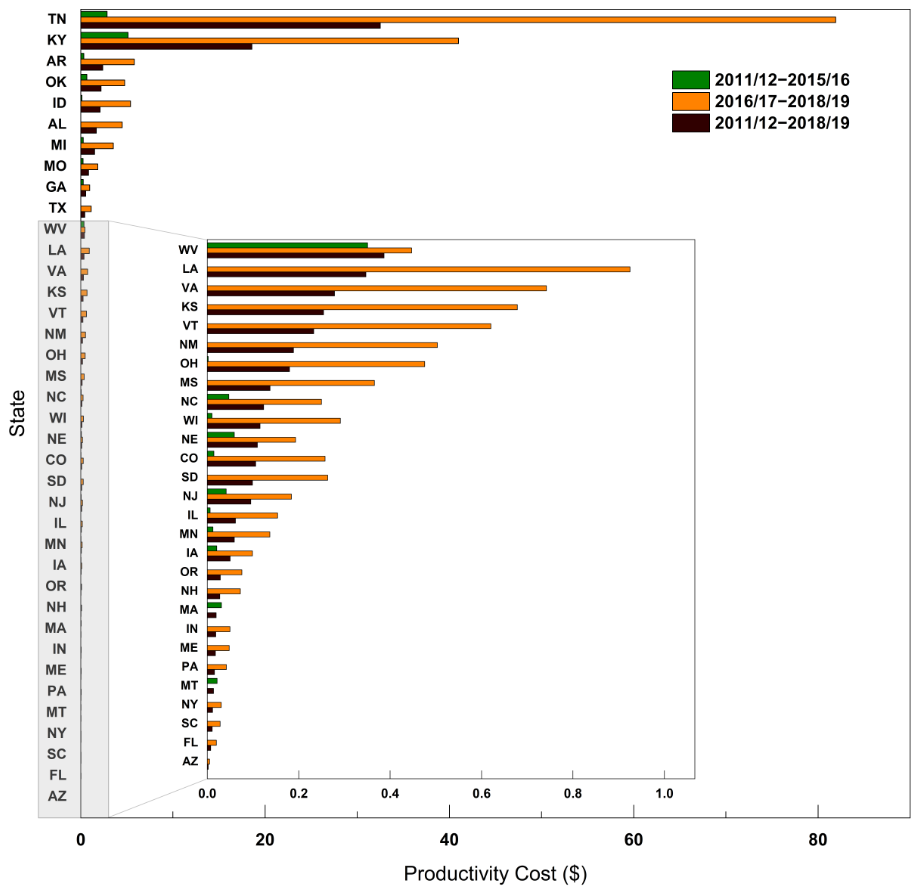

* No ILI-related reactive school closures were observed in public schools in the states of WY,WA, UT,RI, ND, NV, MD, HI, DC, DE, CT, CA, and AK from 2011‒12 to 2018‒19.

ILI, influenza or influenza-like illness
